# Breastfeeding infants receive neutralizing antibodies and cytokines from mothers immunized with a COVID-19 mRNA vaccine

**DOI:** 10.1101/2021.10.12.21264890

**Authors:** Vignesh Narayanaswamy, Brian Pentecost, Corina N. Schoen, Dominique Alfandari, Sallie S. Schneider, Ryan Baker, Kathleen F. Arcaro

## Abstract

**Objective:** To evaluate the immune response to COVID-19 mRNA-based vaccines present in breastmilk and the transfer of the immune response to the breastfeeding child.

**Methods:** A prospective cohort study enrolled 30 lactating women who received an mRNA-based COVID-19 vaccine between January and April 2021. Women provided serial milk samples, which included milk expressed before vaccination, milk expressed across 2-3 weeks after the first dose, and milk expressed across 3 weeks after the second dose. Women also were asked to provide their blood, spotted on cards (dried blood spots; DBS) 19 days after the first dose and 21 days after the second dose. Stool samples from the breastfed infants were collected 21 days after mothers received their second dose. Pre-pandemic samples of milk, DBS cards, and infant stool from prior studies were also utilized. Milk and infant stool samples were tested by ELISA for receptor-binding domain (RBD)-specific IgA and IgG. Milk samples were tested for the presence of neutralizing antibodies against the spike and four variants of concern (VOCs): D614G, B.1.1.7 (alpha), B.1.351 (beta), and P.1 (gamma). Milk samples were also tested by electrochemiluminescence assay for levels of 10 cytokines.

**Results:** Milk from COVID-19-immunized women neutralized the spike and four VOCs and this response is primarily IgG-driven. The immune response in milk also included significantly elevated levels of interferon-γ (IFN-γ). The immune response to maternal vaccination was reflected in breastfed babies; anti-RBD IgG and anti-RBD IgA was detected in 33% and 30% of infant stool samples, respectively. Levels of anti-RBD antibodies in infant stool correlated with maternal vaccine side-effects.

**Conclusion:** Humoral and cellular immune responses to mRNA-based COVID-19 vaccination are present in the breastmilk of most women. The milk anti-RBD antibodies can neutralize SARS-CoV-2 spike and VOCs. Importantly, we describe for the first time the transfer of anti-RBD antibodies to breastfed infants, with the potential to confer passive immunity against SARS-CoV-2.

## INTRODUCTION

Two COVID-19 mRNA-based vaccines, the BNT162b2 Pfizer/BioNTech and mRNA-1273 Moderna/NIH, are approved for use in the U.S^1,2^. The decision on whether to receive COVID-19 vaccines is of concern among women who are breastfeeding because of the lack of sufficient data on both the safety in breastfeeding women and the potential consequences for infants. Clinical trials for the two mRNA vaccine candidates did not include breastfeeding women, but the Centers for Disease Control and Prevention, the American College of Obstetricians and Gynecologists, and the Academy of Breastfeeding Medicine recommend that breastfeeding women receive the vaccine^3,4^. Passive immunity conferred to breastfeeding infants by vaccinated mothers, or to the developing fetus via the placenta of vaccinated pregnant women, are likely the only means of protection for infants from SARS-CoV-2 infections.

New genetic variants of concern (VOCs) from the original SARS-CoV-2 sequence have emerged in the last several months. The D614G variant is associated with increased infectivity^5^, the U.K variant alpha (B.1.1.7) is associated with enhanced transmissibility^6^, and some reports suggest that the South African and Brazil variants (B.1.351; beta and P.1; gamma) evade the natural immunity conferred by a prior SARS-CoV-2 infection^7,8^.

As of September 28, 2021, we identified ten published studies on the immune response to an mRNA-based COVID-19 vaccine in breastmilk of lactating women^9–18^. The number of lactating women included in eight of these studies ranged between 5 and 31; two remaining studies included 84 and 110 lactating women^17,18^. All studies measured levels of SARS-CoV-2-specific IgA, IgG, and in some cases SARS-CoV-2-specific IgM. However, only one study assessed neutralizing ability of antibodies in milk to the spike complex^11^. Importantly, none of these studies measured cellular responses to the COVID-19 vaccine in milk and presence of anti-SARS-CoV-2-specific antibodies in breastfed infants.

We assessed levels of anti-RBD IgA and IgG in serial milk samples and neutralizing capacity against the wildtype spike and four SARS-CoV-2 variants in pre- and post-vaccination milk samples. We also assessed the vaccinated mothers’ milk for changes in levels of 10 cytokines and the infant stool for levels of anti-RBD IgA and IgG.

## METHODS

### Recruitment of Participants

Study details were promoted on the breastmilkresearch.org/ website. Individuals from across the continental U.S. could enroll if they were lactating and scheduled to receive the vaccine or were recently vaccinated with either the mRNA-based Pfizer/BioNTech or Moderna/NIH vaccine. A total of 30 women were enrolled in the study and provided consent under UMass Amherst IRB-approved protocol #2534. Women were asked to complete study questionnaires assessing their general health, lactation status, previous SARS-CoV-2 infection and related symptoms, type of COVID-19 vaccine received, and the timing of COVID-19 vaccination and related maternal and infant side effects. Information was logged via Research Electronic Data Capture (REDCap).

A pre-pandemic set of milk samples was established from 12 women who donated milk between January 2018 and September 2019 as part of a different study (UMass Amherst IRB-approved protocol #1097). A pre-pandemic set of DBS cards was established from 8 reproductive-age women who provided blood between January 2017 and August 2019 as part of a separate study (Baystate Medical Center IRB-approved protocol #568088). Finally, a pre-pandemic set of stool samples was established from 6 infants collected during August 2019 as part of a separate study (UMass Amherst IRB-approved protocol #944). Pre-pandemic milk, maternal DBS, and infant stool samples were collected using the same methods as samples in the vaccine cohort (see *Sample collection*).

### Sample collection

Consented participants were sent kits with instructions for sample collection, storage, and return. Women were asked to provide serial bilateral breastmilk samples at 13 (Pfizer) or 15 (Moderna) timepoints (∼ every 3 days) over 42 (Pfizer) or 48 (Moderna) days. The timepoints of milk donation varied for each woman (**Table S1**). They were instructed to freeze their milk after expression at each timepoint until all samples were collected and were ready to be shipped to UMass Amherst. Milk samples on two consecutive days before receiving the first vaccine dose were requested from women. In addition to milk samples, women also were asked to provide a sample of their blood at two timepoints, 19 days after receiving the first dose, and 21 days after receiving the second dose. Women who consented were asked to collect blood samples on spot cards (Dried Blood Spots; DBS) (Whatman^®^ FTA^®^ card, Millipore Sigma, #WHAWB120205) which were left to dry at room temperature (RT). Consenting women also provided their infants’ stool samples collected 21 days after the mothers received their second dose. Infant stool samples were collected in stool collection tubes (Fisher Sci., Cat No. NC0705093) containing 8 mL of 95% ethanol. After all samples were collected, participants packaged the samples with ice packs in the kit provided and shipped them to the University of Massachusetts using an overnight express courier.

### Processing breastmilk samples to obtain a whey fraction

Equal volumes of bilateral milk samples were mixed to generate a combined sample. Briefly, 500 μL of combined milk was centrifuged at 820 g for 8 minutes. The whey fraction was carefully transferred to a 48-well plate, and samples from the plate were used for the detection of SARS-CoV-2 RBD-specific immunoglobulins and cytokines, and in the neutralization assay.

### Processing DBS to obtain bloodspot eluate

Discs (6 mm diameter) prepared from spot cards were transferred to a 24-well plate. Five hundred microliters of Tris-buffered saline with 0.05% Tween 20^®^ (TBST) were added to the DBS discs and the plate was incubated with gentle shaking overnight at 4°C. Samples of bloodspot eluates were used for the detection of SARS-CoV-2 RBD-specific immunoglobulins.

### Processing infant stool samples

Infant stool samples were received in stool collection tubes containing 95% ethanol. The tube containing stool was vortexed for 20 minutes until a homogenous suspension was achieved. Aliquots of the stool samples were prepared and stored at −20°C. A single aliquot was retrieved at the time of analysis and centrifuged at 4000 g for 20 minutes at 4°C. After centrifugation, the ethanol supernatant was aspirated, leaving behind the stool pellet, to which TBST was added. The tube was vortexed for 5 minutes, centrifuged at 4000 g for 20 minutes at 4°C and the TBST supernatant was used for the detection of total and SARS-CoV-2 RBD-specific immunoglobulins.

### Enzyme-Linked Immunosorbent Assay (ELISA) for detection of total and SARS-CoV-2-specific immunoglobulins

Levels of SARS-CoV-2 RBD-specific immunoglobulins were measured as previously described^19^. For detection of total immunoglobulins, 96-well plates were coated with anti-IgA (α-chain specific) or anti-IgG (H+L specific) capture antibodies at 1 μg/mL (Jackson ImmunoResearch Laboratories, Inc.). Subsequent steps followed the protocol for detection of RBD-specific immunoglobulins^19^.

### Antibody Neutralization Assay

The neutralization assay was performed using a V-PLEX SARS-CoV-2 Panel 6 multiplex assay by Mesoscale Discovery (K15436U, MSD, Gaithersburg, MD). The assay quantitatively measures antibodies in the sample that can inhibit the interaction of spike and its variants with ACE2. Each plate included an 8-point standard curve. We are reporting results for wildtype spike (referred to as spike), and four spike variants: D614G, B.1.1.7 (alpha), B.1.351 (beta), and P.1 (gamma). All samples and standards were run in technical duplicates. We measured neutralizing ability in milk from the 28 of the 30 women who provided samples both prior to the first dose and after the second dose. For the second time point, the milk sample with the highest level of anti-RBD IgG was selected for each woman, typically the final, penultimate or antepenultimate-collected sample (**Appendix 1**).

### Cytokine Assay

We measured cytokines in milk from the 26 of the 30 women who completed the questionnaire sections on side-effects. Milk samples were selected to represent three time points: prior to vaccination, after the first vaccine dose, and after the second dose. For participants who reported side-effects, the milk samples associated with the first day of reported side-effects after each of the first and second doses were selected (**Appendix 1**). For participants who reported no side-effects, the samples one day after the first and second doses were selected. Cytokines were measured in a multiplex assay (MSD, Gaithersburg, MD) according to the manufacturer’s instructions using 10-plex human V-PLEX Proinflammatory Panel 1 plates (K15049D). Each 96-well plate included an 8-point standard curve and assays for 10 cytokines: IL-2, IL-4, IL-6, IL-8, IL-10, IL-12p70, IL-13, IL-1β, IFN-γ, and TNF-α. All samples and standards were run in technical duplicates.

### Statistical Analysis

Participant characteristics with continuous outcome measures are reported as mean and range, whereas characteristics with categorical outcomes are reported as percentages. The thresholds for positivity for RBD-specific antibodies were set at OD values 3 times above the standard deviation of the OD values obtained with only the secondary antibody (background). For the neutralization assay, percent inhibition was computed using an equation provided by the manufacturer, which is a measure of antibodies that can inhibit the binding of ACE2 to the spike or its variants. Matched paired *t*-tests were used to analyze differences between percent inhibition of neutralizing antibodies in milk provided before vaccination and milk provided after the second dose. Matched paired *t*-tests were used to analyze differences in cytokine levels between the indicated timepoints. Independent *t*-tests were used to analyze differences in infant stool anti-RBD antibodies, stratified by the presence of maternal side effects to immunization. Pearson R was used to determine the correlation between anti-RBD IgG levels and percent inhibition of SARS-CoV-2 antigens (neutralization). *P* values < 0.05 were considered statistically significant. Statistical analyses were performed using GraphPad Prism 9.

## RESULTS

### Participant demographics and vaccination details

Demographic characteristics and COVID-19 vaccination-related information are summarized in **Table 1**. Thirty lactating women who were scheduled to receive or received an mRNA-based COVID-19 vaccine were enrolled. The study population consisted of women who self-identified as white (*n* = 27), black (*n* = 1), or Asian (*n* = 2) and their breastfeeding infants. At the time of enrollment, women were between 26 to 46 years of age and their infants ranged from 7 days to 21.7 months old. Twenty study participants received the Pfizer vaccine and 10 received the Moderna vaccine. Three of 30 women reported a prior positive test for SARS-CoV-2 with a known timing of infection and symptoms. Serial milk samples were obtained from all 30 women; two of thirty women did not provide milk samples prior to receiving their first vaccine dose and one woman provided serial milk samples only after receiving her second dose. Twenty-eight women provided DBS samples, of whom 20 provided DBS samples at both timepoints. Infant stool samples were received from 24 of the 30 vaccinated participants.

**Table 1.** Participant demographics and vaccination-related information (*n* = 30)

| Characteristics | Mean (Range) |  |
| --- | --- | --- |
| Mother's Age<sup>a,b</sup> (year) | 35 (26-46) |  |
| Baby's Age<sup>a,b</sup> (Days) | 239 (7-651) |  |
| BMI<sup>a</sup> (kg/m<sup>2</sup>) | 27 (20-39) |  |
|  | ***n* (%)** |  |
| **Race** |  |  |
| White | 27 (90) |  |
| Black | 1 (3) |  |
| Asian | 2 (7) |  |
| **Previous SARS-CoV-2 infection** | 3 (10) |  |
| **Vaccine type** |  |  |
| Pfizer/BioNTech | 20 (67) |  |
| Moderna | 10 (33) |  |
| **Number of women who provided milk** |  |  |
| Before first dose | 28 (93) |  |
| After first dose | 29 (97) |  |
| After second dose | 30 (100) |  |
| **Number of women who provided DBS cards** |  |  |
| After first dose<sup>c</sup> and after second dose<sup>d</sup> | 20 (67) |  |
| Only after first or after second dose provided | 8 (27) |  |
| None provided | 2 (7) |  |
| **Infant stool<sup>e</sup> samples provided** | 24 (80) |  |
| **Maternal side effects** | **After first dose *n* (%)** | **After second dose *n* (%)** |
| Fever symptoms | 4 (13) | 8 (27) |
| Tiredness | 4 (13) | 11 (37) |
| Aches | 5 (17) | 8 (27) |
| Runny nose | 0 (0) | 1 (3) |
| Sore throat | 0 (0) | 2 (7) |
| Headache | 4 (13) | 9 (30) |
| No side effect | 18 (60) | 10 (33) |
| Questionnaire not completed | 4 (13) | 3 (10) |
| **Infant side effects** | **After first dose *n* (%)** | **After second dose *n* (%)** |
| Fever symptoms | 2 (7) | 0 (0) |
| Runny nose | 1 (3) | 2 (7) |
| No side effect | 22 (73) | 22 (73) |
| Questionnaire not filled | 5 (17) | 5 (17) |

<sup>a</sup>One woman did not provide information on her age, her baby's age and BMI; <sup>b</sup>Age reported at the time of enrollment; <sup>c</sup>DBS eluates provided 19-23 days after the first dose; <sup>d</sup>DBS eluates provided 19-23 days after the second dose; <sup>e</sup>Infant stool samples provided 21 days after the second dose.

Side effects after receiving the first and second dose of both vaccine types are summarized in **Table 1**. Overall, the number of women who reported experiencing any side effects was greater after they received the second dose as compared to after the first dose. The number of women who reported vaccine-related fevers (27%), aches (27%), tiredness (37%), or headaches (30%) was greater after the second dose. Most mothers (73%) reported that their infants had no side-effects. When infant side-effects did occur, they included fever and/or runny nose (**Table 1)**.

### Functional antibodies are detected in breastmilk and serum from vaccinated women

We measured RBD-specific IgA, IgG, and IgM in serial milk samples provided by the 30 vaccinated participants. Overall, IgM levels were consistently negligible, and data are not shown. Milk samples from 26 of the 30 women provided more than 2 weeks after the second dose were positive for RBD-reactive IgG, albeit with a dynamic range (**Figures 1A and B**). RBD-reactive IgG levels increased two to four-fold above the positive cut-off value in the milk from 3 of the 20 women who received the Pfizer vaccine, after they received the first dose (**Figure 1A**). However, among women in the Moderna cohort, RBD-reactive IgG levels only exceeded the assay’s positive cut-off limit after their second dose (**Figure 1B**). In contrast to IgG responses, milk samples from only 14 of 30 women were positive for RBD-reactive IgA (**Figures 1C, D, and Figure 2A**). Lactational stage, as measured by baby’s age, was not related to the immune response in milk; milk from women with infants aged 1.5 months and 23 months had comparable levels of anti-RBD IgA and IgG (**Appendix 2, A and B**)

**Figure 1.**
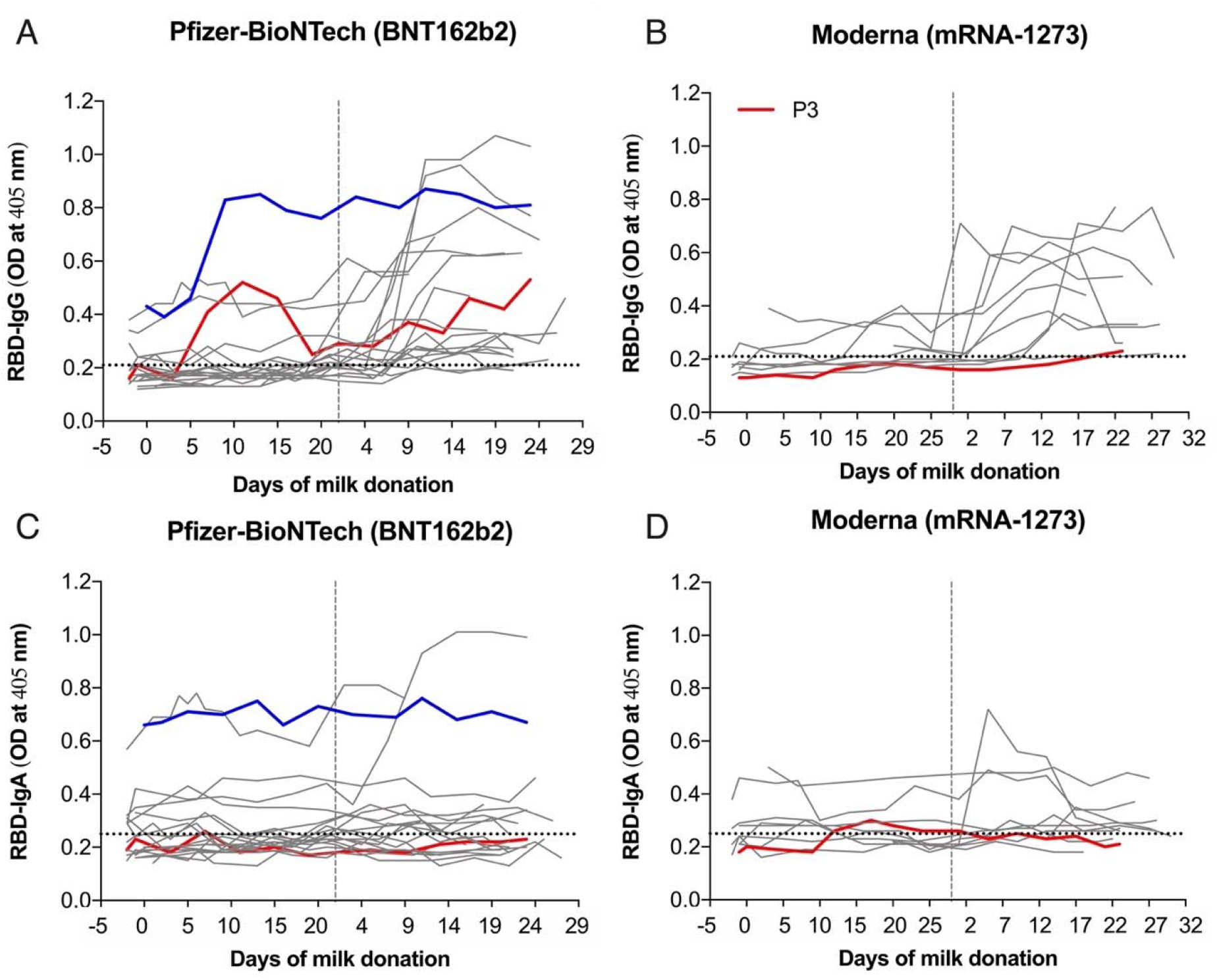
Antibody levels in serial milk samples following COVID-19 mRNA vaccination. Milk samples were obtained prior to the first dose (timepoint 0), across 19-23 days after the first dose and across 19-23 days after the second dose (vertical dashed line) from 30 women vaccinated against SARS-CoV-2. Whey fractions were assessed with ELISA for RBD-specific IgG (A and B) and IgA (C and D). Colored lines indicate serial milk samples obtained from 3 women who had a previous positive diagnosis for COVID-19. Horizontal dotted lines indicate the positive cut-off values.

**Figure 2.**
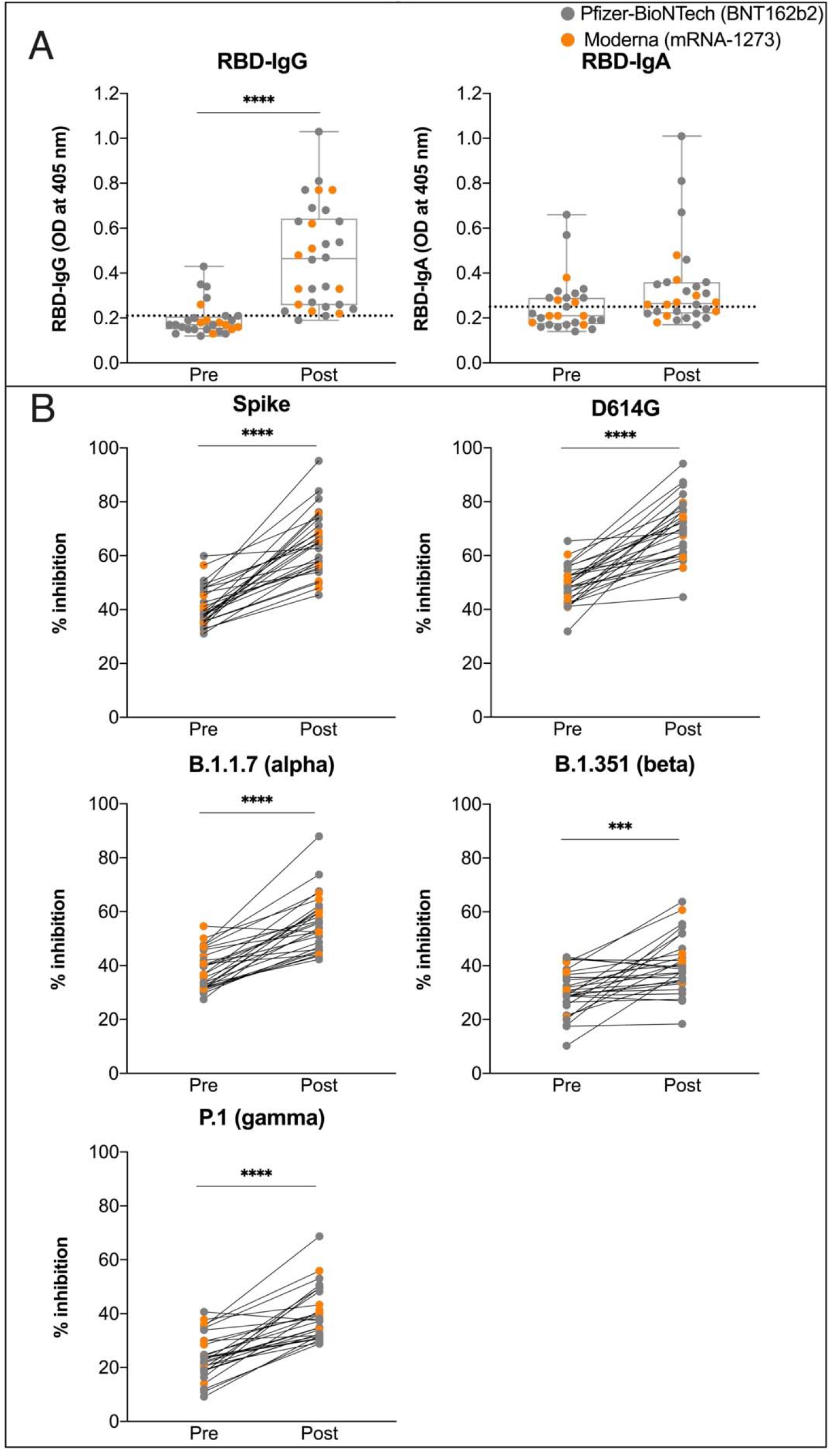
Milk samples from vaccinated women have neutralizing capacity against SARS-CoV-2 spike complex. Comparison of levels of RBD-specific IgG and IgA (A) in milk provided prior to vaccination (Pre) and 3 weeks after the second dose (Post). Dotted horizontal lines indicate positive cut-off values. (B) Comparison of percent inhibition of spike and its variants of concern (D614G, B.1.1.7: alpha, B.1.351: beta, and P.1: gamma) between milk provided prior to vaccination (Pre) and 3 weeks after the second dose (Post). Orange data points indicate Pfizer vaccine recipients; Grey data points indicate Moderna vaccine recipients. Matched paired *t*-tests were performed to analyze differences in percent inhibition between Pre and Post. \*\*\*\**p* < 0.0001 \*\*\**p* = 0.0001

Among the 30 vaccinated women, three reported that they had a prior COVID-19 positive test result. Two of the 3 women received the Pfizer vaccine (P1 and P2; **Figure 1A and C; blue and red lines**) and one woman received the Moderna vaccine (P3; **Figure 1B and D; red line**). Time from positive diagnosis to receiving the first vaccine dose was 210, 77, and 56 days for P1, P2, and P3, respectively. Of the three women, only P1 had high a level of anti-RBD IgA in milk obtained before vaccination (before day 0), and these levels remained consistent across all time points of milk donation (**Figure 1C; blue line**). Unlike anti-RBD IgA, the level of anti-RBD IgG increased by approximately 2-fold 10 days after participant P1 received her first dose, and then stayed high across later time points (**Figure 1A**). Milk obtained prior to vaccination from participants P2 and P3 was negative for anti-RBD antibodies (**Figures 1A-D**), despite positive COVID-19 tests 77 days (P2) and 56 days (P3) before initial vaccination.

Breastmilk and serum humoral response to the vaccines were primarily IgG driven. Serum RBD-specific IgA levels were at or below background in DBS eluates from all 20 women (*data not shown*). Eight of the 30 women provided DBS cards at only one time-point and the remaining two women did not provide DBS samples (**Table 1**). Similarly, median RBD-specific IgA did not increase in milk from women after the second dose compared to pre-vaccine milk (**Figure 2A**). In contrast, median RBD-specific IgG increased in milk from women after the second dose (**Figure 2A**), along with an increase in median anti-RBD IgG in serum (**Appendix 3**).

We assessed RBD-specific antibodies in a set of 12 pre-pandemic milk samples (collected during January 2018 – September 2019). Median levels of milk RBD-IgA and RBD-IgG were below positive cut-off values (**Appendix 4**). However, 3 samples scored positive for anti-RBD-IgA, and 2 samples scored positive for anti-RBD-IgG.

To determine functionality, we evaluated the presence of vaccine-elicited neutralizing antibodies in milk samples of 28 women; two of the 30 women did not provide milk prior to the first dose (**Table 1**). The milk samples from before vaccination and the milk samples that had the highest RBD antibody levels were assayed for each woman. Milk neutralizing antibodies to spike and four VOCs (D614G, alpha, beta, and gamma) were evaluated^20^. Antibody neutralization is reported as percent inhibition of binding by purified ACE2 to immobilized targets (spike or its VOCs). Analysis of paired samples shows that milk samples provided more than 3 weeks after the second dose were able to inhibit the binding of ACE2 to the spike (*p* < 0.0001) (**Figure 2B**). Similarly, these milk samples inhibited the binding of ACE2 to four VOCs; however, the ability to inhibit the binding of ACE2 to the beta variant (B.1.351) was more limited (**Figure 2B**). The neutralizing ability of milk for spike correlated with levels of RBD-specific IgG (Pearson’s R = 0.36; *p* = 0.03) (**Figure 3**). This also was observed for 3 of the 4 VOCs; D614G (*p* = 0.04), alpha (*p* = 0.01), and gamma (*p* = 0.04) (**Appendix 5**). As with anti-RBD-IgA and anti-RBD-IgG responses, there was no relationship between neutralizing ability and lactation stage (baby’s age) (**Appendix 2, C**).

**Figure 3.**
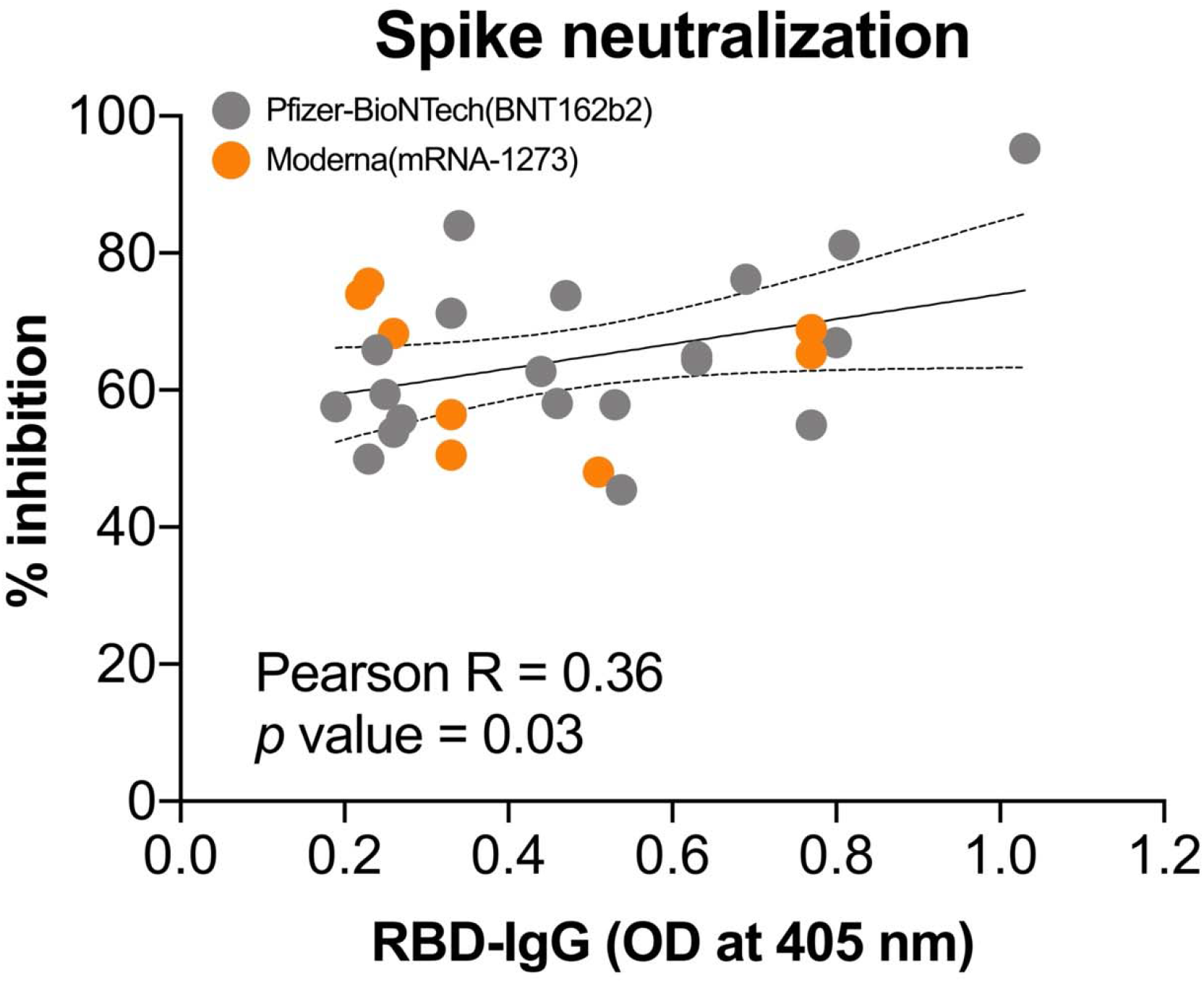
Correlation between anti-RBD IgG levels and percent inhibition of SARS-CoV-2 spike binding to ACE2. Neutralization ability was assessed in milk collected 21-23 days after the second dose using the MSD V-PLEX SARS-CoV-2 Panel 6 assay. The correlation between anti-RBD IgG and neutralization ability determined in the milk of 28 women. 95% confidence interval, Pearson’s correlation coefficient (R) and *p* value are shown.

### SARS-CoV-2-reactive antibodies were detected in stool obtained from infants of vaccinated mothers

We first measured levels of total IgA and IgG to validate our ELISA protocol on ethanol-preserved stool samples (*see Methods*). Stool samples from 24 infants were assessed; median total IgA and IgG levels were above positive cut-off limits (**Appendix 6**). Detection of total IgA and IgG indicates that the ELISA was optimized for stool samples collected in 95% ethanol. Next, we assessed levels of anti-RBD IgA and IgG. We detected anti-RBD IgA and IgG in 30% and 33% of infant stool samples, respectively (**Figures 4A and B**); these infants ranged in age from 55 days to 11 months. We found that levels of anti-RBD IgG antibodies in infant stool were higher when mothers had experienced side effects to the vaccine (**Figures 4C and D**). Pre-pandemic stool samples (*n* = 6) contained IgA and IgG (**Appendix 6)** but were negative for anti-RBD antibodies (**Appendix 4**).

**Figure 4.**
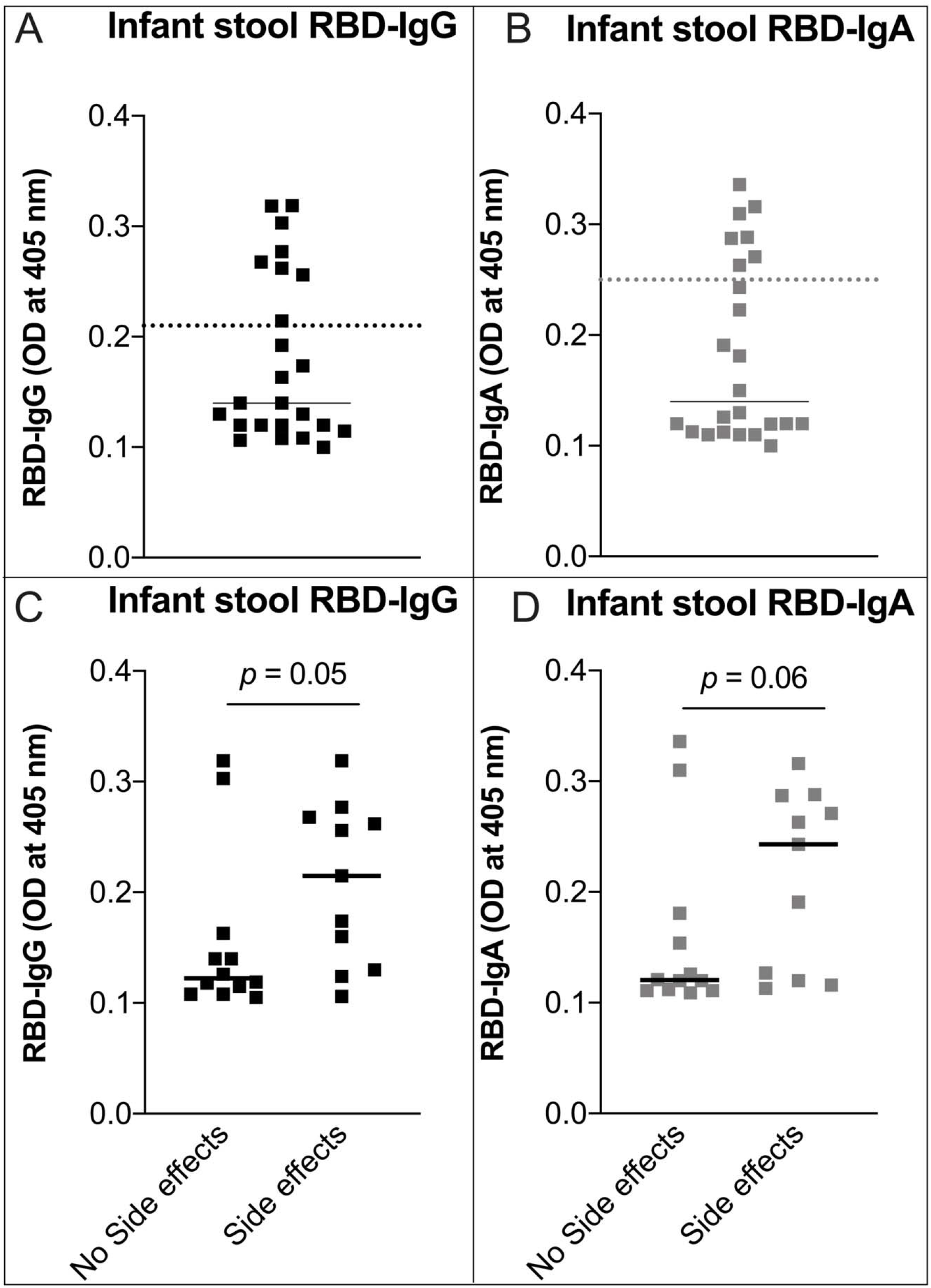
Anti-RBD IgG and IgA levels in infant stool are higher in infants of mothers who reported side effects after vaccination. Levels of anti-RBD IgG (A) and anti-RBD IgA (B) in stool samples obtained from infants (*n* = 24) of vaccinated mothers. Horizontal dotted lines indicate positive cut off values. Comparison of levels of anti-RBD IgG (C) and anti-RBD IgA (D) between stool obtained from mothers who reported no side effects and those who reported vaccine-related side effects. Differences in levels of infant stool anti-RBD antibodies stratified by the presence of maternal side effects were assessed with independent *t*-tests.

### Cell-mediated responses were detected in breastmilk from vaccinated women

We measured levels of ten key cytokines in milk from 26 vaccinated lactating women; four of 30 women did not complete the questionnaire on side effects (**Table 1**). Cytokines included IL-2, IL-4, IL-6, IL-8, IL-10, IL-12p70, IL-13, IL-1β, IFN-γ, and TNF-α. All analytes, except IL-12p70 and IL-4, were detectable in all samples. Data for IL-12p70 and IL-4 are not discussed further. The levels of IFN-γ were significantly higher in milk provided following the first dose and after the second dose as compared to milk provided before receiving the vaccine (*p* < 0.05; *p* < 0.01, respectively, **Figure 5A**). Data were next analyzed based on whether women experienced side-effects to the vaccine. Among the women who reported no side effects after either the first or second dose (*n* = 13), compared to samples provided prior vaccination, the median levels of IFN-γ levels increased by approximately 2-fold in samples provided following the first dose, and by 3-fold in samples provided following the second dose (**Figure 5B**). Among the women who reported any side effects (*n* = 13), compared to samples provided before vaccination, the median levels of IFN-γ increased by approximately 2.5-fold in samples provided following the first dose, and by over 20-fold in samples provided following the second dose. Overall, among the women who reported any side effects, the levels of IFN-γ were significantly higher in milk provided following the second dose compared to milk provided before receiving the vaccine (*p* < 0.01) (**Figure 5B**). Levels of the remaining seven cytokines were comparable across the three timepoints (**Appendix 7**).

**Figure 5.**
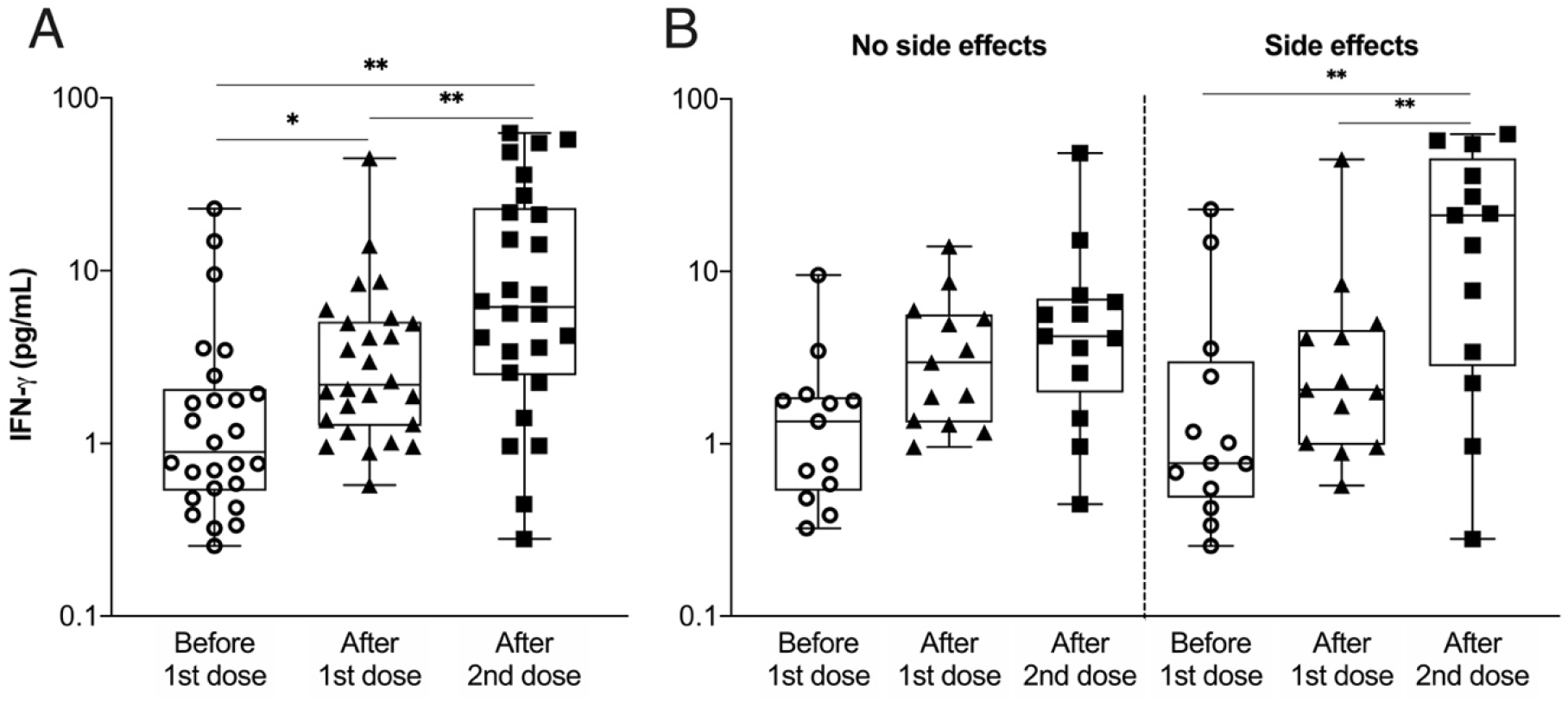
IFN-γ levels increase in milk after women are vaccinated. (A) IFN-γ levels were assessed in milk obtained from women (*n* = 26) at three timepoints: prior to COVID-19 vaccination, following first dose, and following the second dose. (B) IFN-γ levels assessed in milk at the three timepoints after stratifying women by whether they reported no side effects, or side effects after either the first or second dose. Horizontal lines in each box indicate median concentration (pg/mL). \**p* < 0.05; \*\**p* < 0.01

## DISCUSSION

Thus far, we identified ten published studies on the immune response to an mRNA-based COVID-19 vaccine in breastmilk of lactating women^9–18^. Our study adds new information on the vaccine-induced immune response: we show neutralizing ability of antibodies in milk against spike and 4 VOCs that is IgG-driven and high levels of IFN-γ in milk. Additionally, we show for the first time the presence of anti-RBD antibodies in the stool of breastfed infants.

Breastmilk from vaccinated women contained elevated levels of antibodies against RBD. Induction of RBD-reactive IgG in milk occurred after the second dose in 87% of lactating, vaccinated women and we also detected RBD-reactive IgG in blood from all women assayed. In contrast to the IgG response in milk, RBD-reactive IgA was detected in only 47% of lactating, vaccinated women; and we did not observe a concomitant increase in anti-RBD IgA in blood. The lack of a robust anti-RBD IgA response in breastmilk and blood of vaccinated women can be attributed to the intramuscular route of administration of the mRNA vaccines^21^. In addition, class switching to IgG occurs very quickly after COVID-19 vaccination^22,23^ in the absence of prior exposure, which explains why we did not detect RBD-reactive IgA in most milk samples. In a disease state where SARS-CoV-2 is mucosally-acquired, we and others find the immune response mounted is primarily IgA^19,24–27^. This is seen for one of the three vaccinated women in our study who also previously tested positive for SARS-CoV-2. She had high levels of anti-RBD IgA in her milk pre-vaccination.

We assessed the ability of antibodies in milk to neutralize spike and four circulating VOCs. Milk provided after women received their second dose had significantly increased neutralizing ability as compared to their matched pre-vaccine milk samples. The neutralization ability of milk was significant against spike and all four VOCs, however, we observed considerable variability among women. The milk of nine women showed no change in neutralization ability against B.1.351 (beta) after the second dose, despite exhibiting neutralizing ability against spike (**Appendix 8**). Studies of post-vaccination serum also report reduced neutralizing antibody titers against the B.1.351 variant as compared to spike^8^ and Collier *et al*^11^ report limited serum and milk antibody titers, from vaccinated lactating and pregnant women, against the B.1.351 variant. The individual differences among the milk of women to selectively neutralize specific variants warrants further study.

In most cases, the side effects of COVID-19 mRNA vaccines include fevers, headache, and/or myalgia. The extent to which these side effects correlate with the vaccine-induced cytokine response has received little attention. We explored this relationship and found elevation of the cytokine IFN-γ in breastmilk. Most side effects can be attributed to the production of type I interferons, which play an important role in enhancing immune response in the early stages following vaccination. Type I interferons act in synergy with IFN-γ (a type II interferon), and both are key cytokines induced after an mRNA vaccination^28^. Interferons induce activation of dendritic cells, enabling these cells to present the translated RBD to naïve CD4^+^ and CD8^+^ T cells, which are part of the adaptive immune system. This is crucial because activated CD4^+^ T cells stimulate B cells to produce RBD-specific antibodies. Furthermore, interferons promote the formation of long-lived memory CD4^+^ and CD8^+^ T cells^29,30^. Thus, a key finding of our study is the elevation of IFN-γ in milk of women receiving an mRNA vaccine. This should protect nursing infants against several viral respiratory tract infections, including SARS-CoV-2^31,32^.

Infants of mothers previously unexposed to SARS-CoV-2 presumably have an underdeveloped immune response to the virus. We assessed stool samples from infants of the mothers immunized with a COVID-19 mRNA vaccine. Prior studies indicate that the infant GI tract does not degrade IgA or IgG^33,34^; we detected total IgA in all infant stool samples and total IgG in all but one stool sample, consistent with prior studies^33,35^. We assessed anti-RBD IgA and IgG in infant stool of vaccinated mothers. Importantly, our findings show detectable levels of anti-RBD IgA and IgG in 30% and 33% of infant stool samples. Detection of these antibodies in infant stool provides compelling evidence for the immunity conferred to infants from vaccinated mothers. It is well known IgA mediates an early neutralizing response at mucosal surfaces^25,36^; therefore, detection of anti-RBD IgA in stool samples in our cohort suggests protection of these infants from a potential SARS-CoV-2 infection not only systemically (contributed by anti-RBD IgG), but also at mucosal sites mediated by anti-RBD IgA.

The potential sources of infant anti-SARS-CoV-2 antibodies are cord blood and breastmilk. The transfer of passive immunity through cord blood depends on women getting the COVID-19 vaccine while pregnant. However, preliminary research shows limited neutralizing potency of anti-RBD IgG transferred through cord blood in SARS-CoV-2 positive^21^ and COVID-19 vaccinated^13^, pregnant women. Additionally, since IgA is not known to cross the placental barrier, the extra protection conferred by anti-RBD IgA from milk is a motivation to continue breastfeeding after receiving a COVID-19 vaccine.

## Data Availability

All data produced in the present work are contained in the manuscript

## Acknowledgements

The authors are grateful to Rachel Taylor for her help with sample collection.

## Author contributions

*Concept and design*: Arcaro, Pentecost, Narayanaswamy, Schoen

*Sample collection*: Arcaro, Pentecost, Narayanaswamy, Schoen, Schneider

*Laboratory experiments*: Narayanaswamy and Baker

*Interpretation of data*: Arcaro, Pentecost, Narayanaswamy

*Drafting of the manuscript*: Narayanaswamy, Arcaro, Pentecost

*Funding*: Arcaro and Alfandari

All authors read and edited earlier versions and approved the final article.

