## Supplementary files for "Breastfeeding infants receive neutralizing antibodies and cytokines from mothers immunized with a COVID-19 mRNA vaccine"

Narayanaswamy, V *et al*

**Appendix 1**. Complete data-set submitted as excel file.

**Appendix 2. No relationship between lactation stage and immune response in milk.** (A and B) Correlation between milk anti-RBD antibodies and lactation stage as measured by baby’s age (months). (C) Correlation between milk neutralization of wildtype spike and lactation stage as measured by baby’s age (months). Pearson’s correlation coefficient (R) and *p*-values are shown.

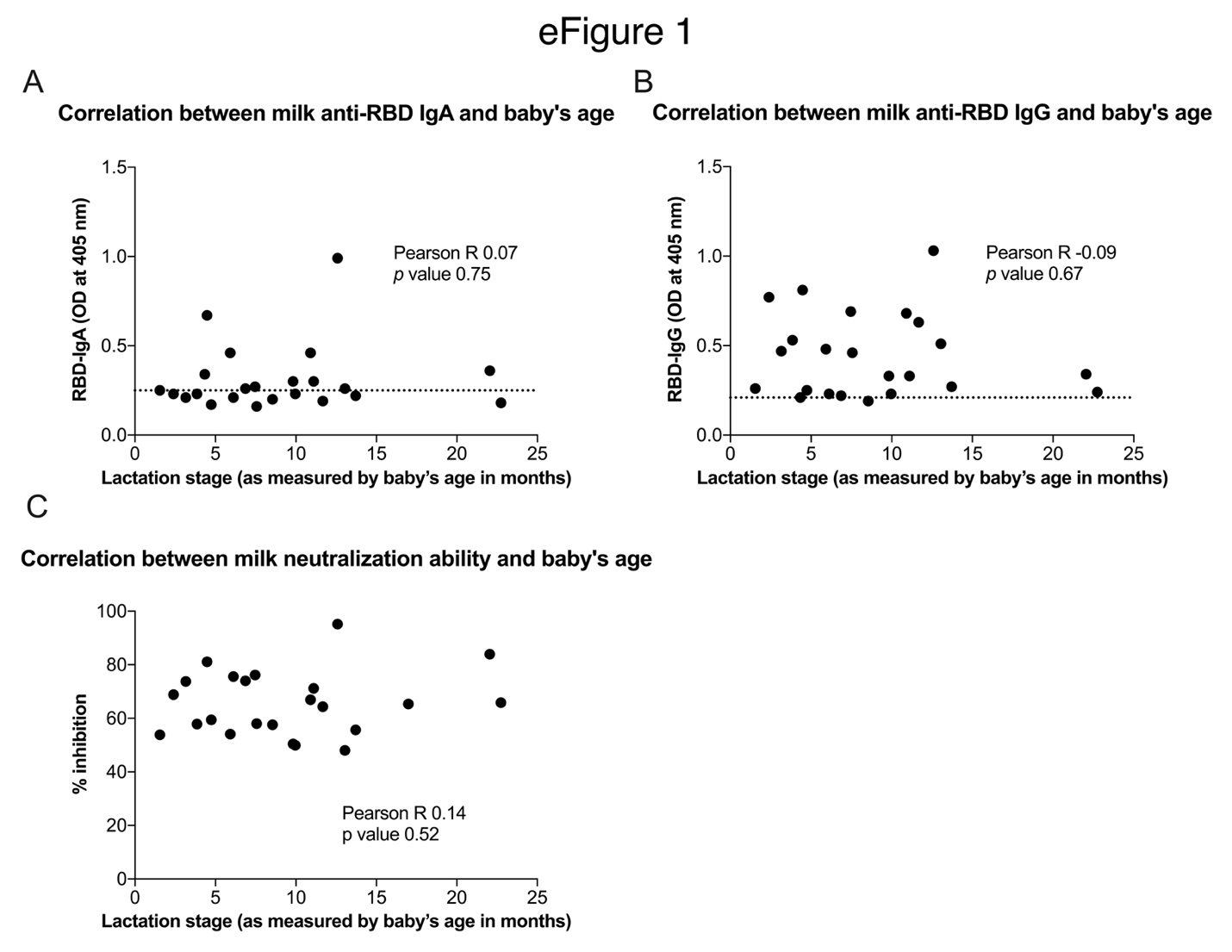

**Appendix 3. Anti-RBD IgG detected in dried blood spots (DBS) 21 days following second dose of vaccination.** Comparison of anti-RBD IgG in DBS eluates provided by women in the pre-pandemic control group (*n* = 8), by women 19 days after the first vaccine dose and 21 days after the second dose (*n* = 21). Differences between DBS first dose and DBS second dose were assessed with a matched paired *t*-test. Differences between DBS second dose and pre-pandemic DBS controls were assessed with an independent *t*-test. *****p* ≤ 0.0001

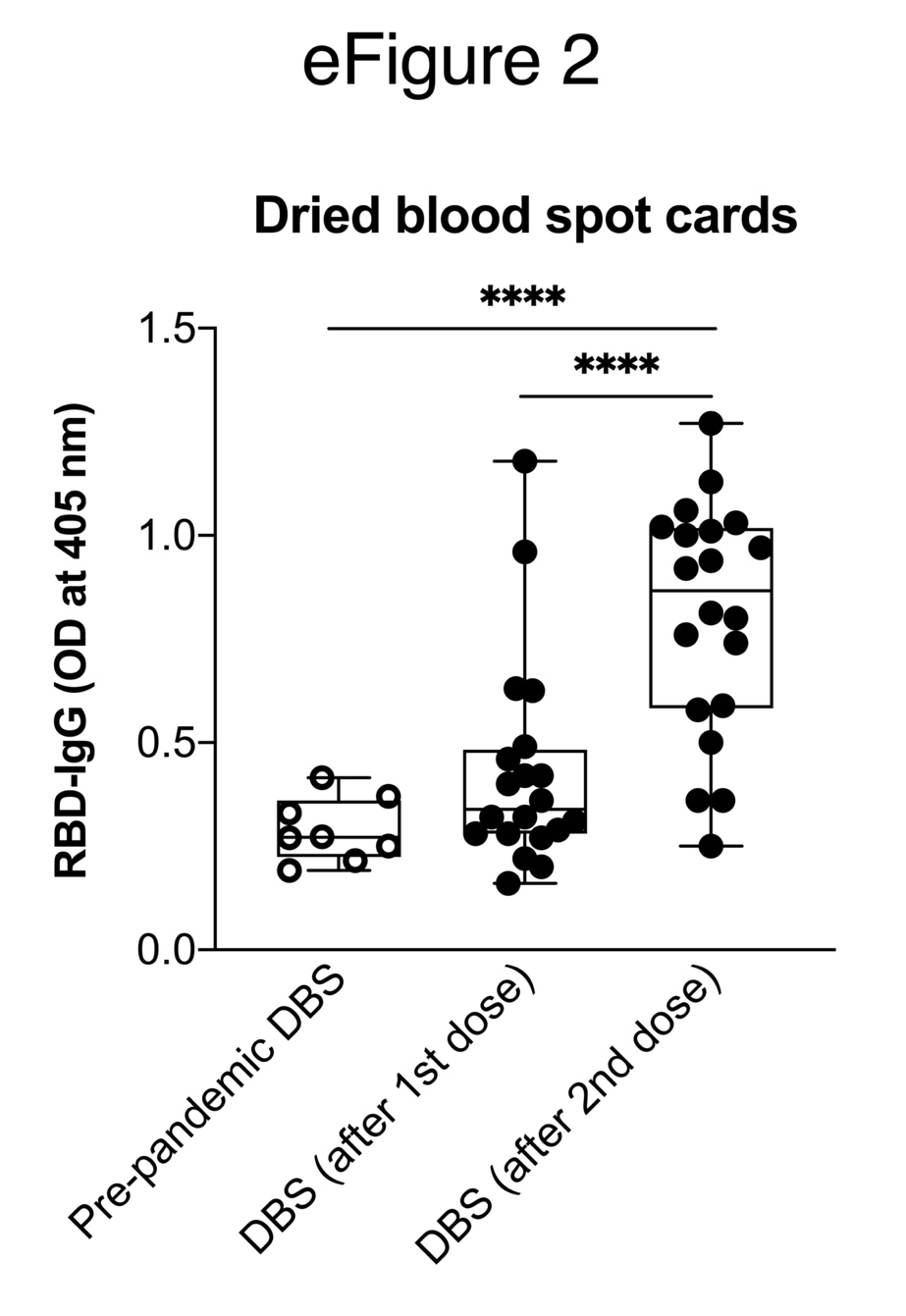

**Appendix 4. Anti-RBD IgA and IgG are not detected in pre-pandemic milk and stool controls.** Levels of milk anti-RBD IgA and IgG (circle; *n* = 12) and levels of infant stool anti-RBD IgA and IgG (square; *n* = 6) detected by ELISA. Dotted lines indicate positive cut-off values for anti-RBD IgA (grey) and anti-RBD IgG (black). Solid lines indicate median ODs.

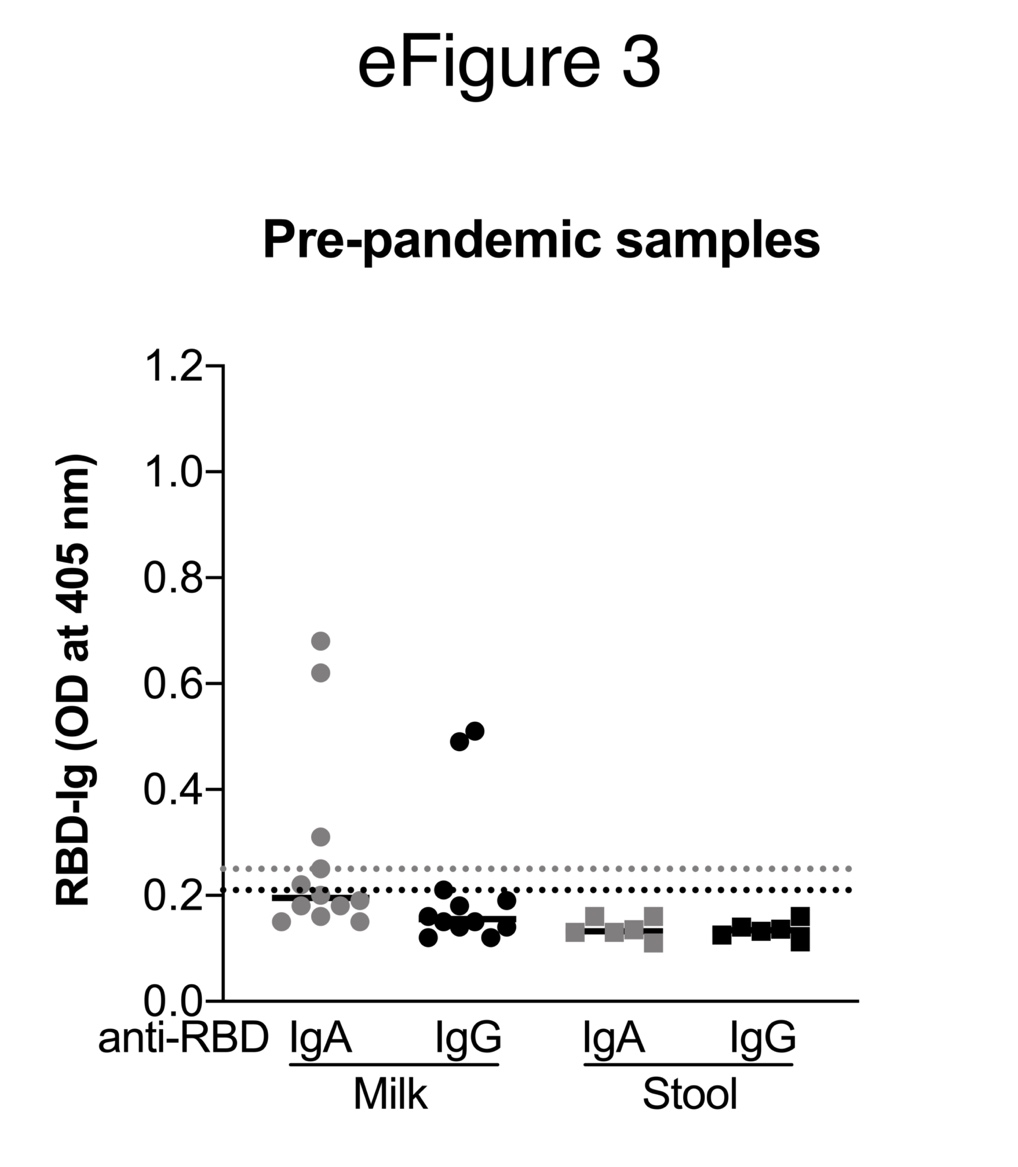

**Appendix 5. Correlation between anti-RBD IgG and percent inhibition of SARS-CoV-2 variants of concern**, including D614G (A), B.1.1.7 (B), B.1.351 (C), and P.1 (D). 95% confidence intervals, Pearson’s correlation coefficient (R) and *p* values are shown.

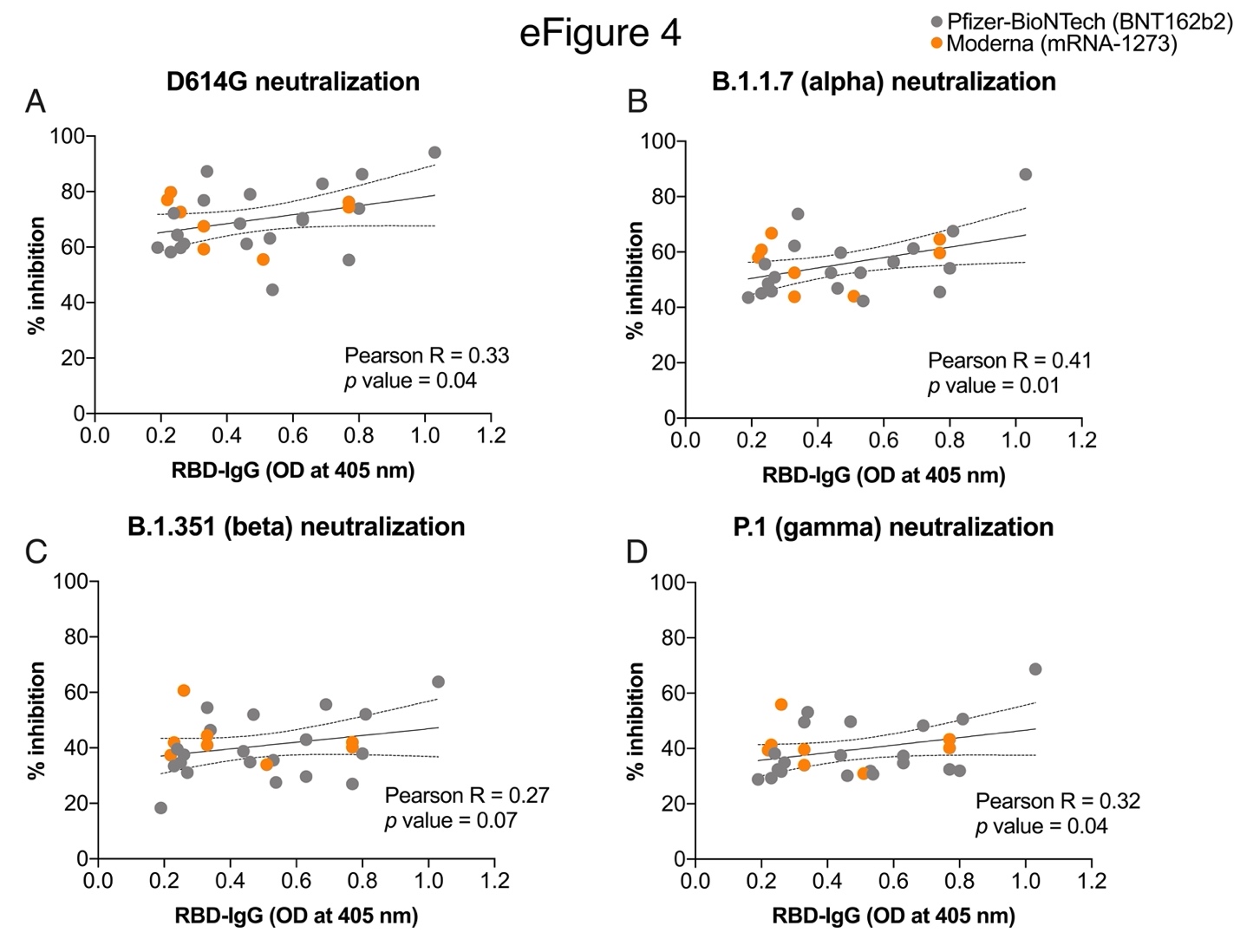

**Appendix 6. Total IgA and IgG are present in infant stool samples in the vaccine cohort and pre-pandemic controls**. Levels of total IgA and IgG in stool from infants of COVID-19 immunized mothers (filled square; *n* = 24) and levels of total IgA and IgG in pre-pandemic infant stool samples (open square; *n* = 6) detected by ELISA. Dotted lines indicate positive cut off values, IgA in gray and IgG in black. Solid lines indicate median ODs.

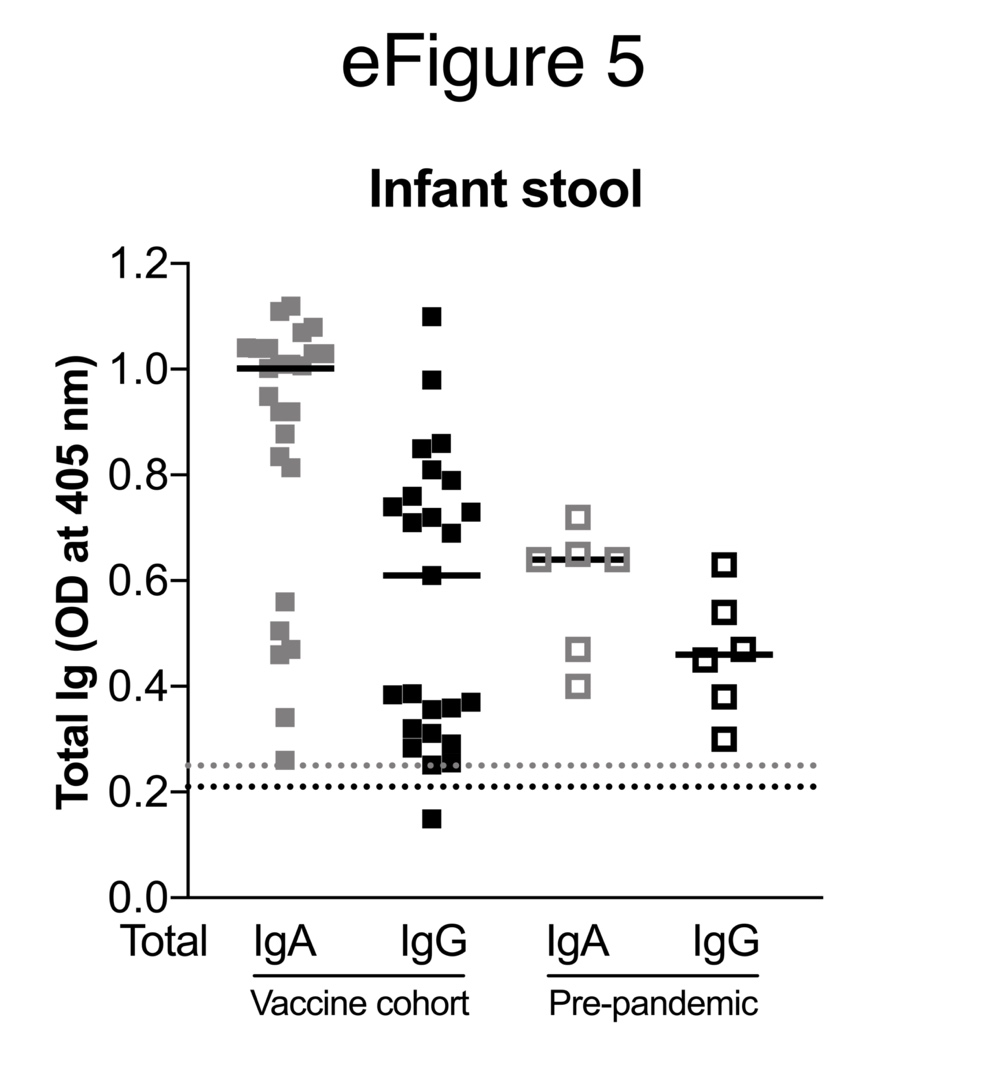

**Appendix 7. No significant change in levels of 7 cytokines assayed in milk obtained before and after vaccination against COVID-19.** Cytokine levels were assessed in milk samples obtained from 26 women prior COVID-19 immunization, after the first dose, and after the second dose. Horizontal lines in each bar indicate median concentration (pg/mL).

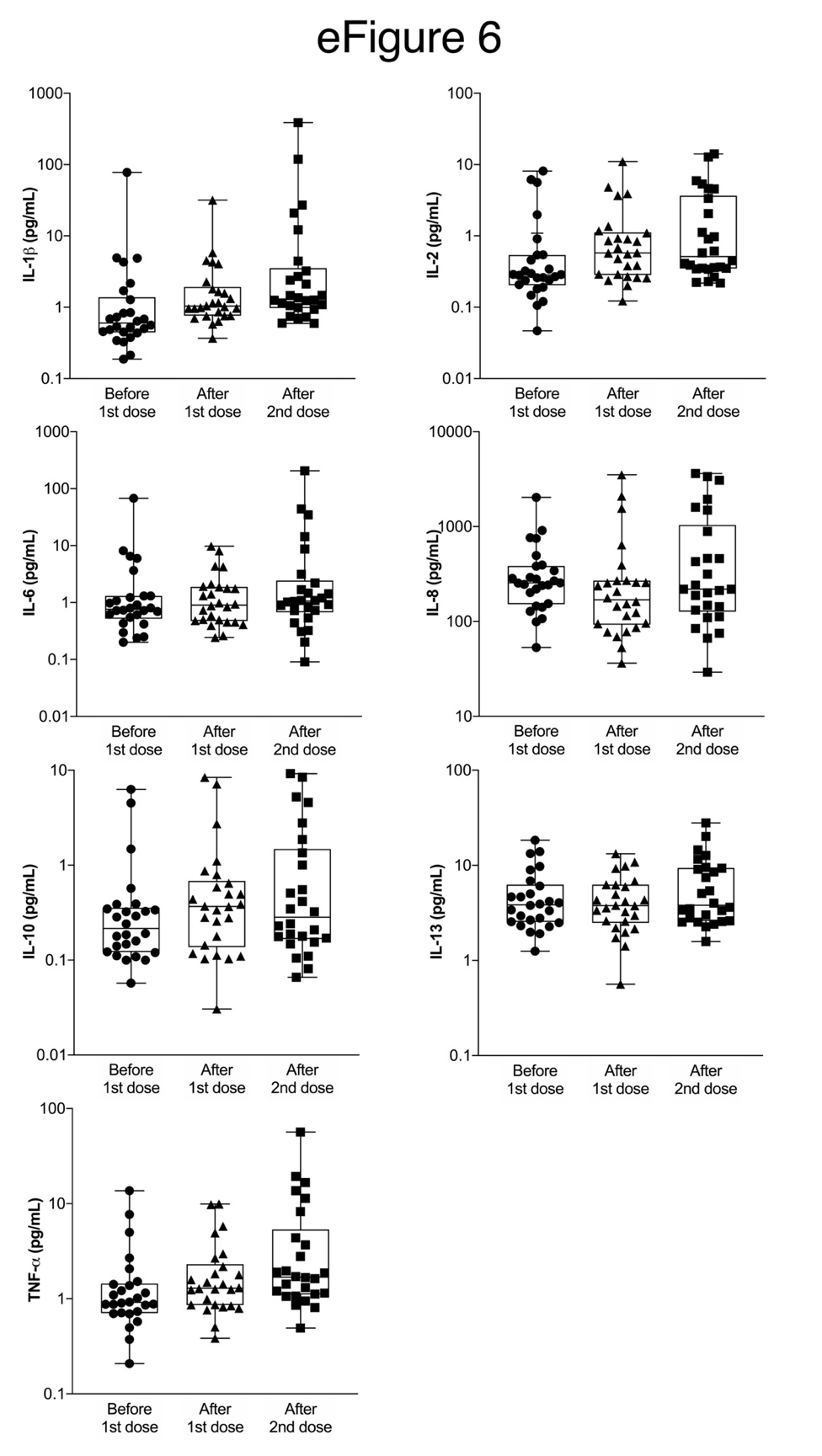

**Apppendix 8. Individual variability in the ability of post-vaccine milk to neutralize spike and the B.1.351 variant**. Milk from nine women who received an mRNA-based COVID-19 vaccine was able to neutralize the spike but not the B.1.351 variant. A matched paired *t*-test was used to compare differences in percent inhibition between pre and post-vaccine milk samples. ****p* < 0.001.

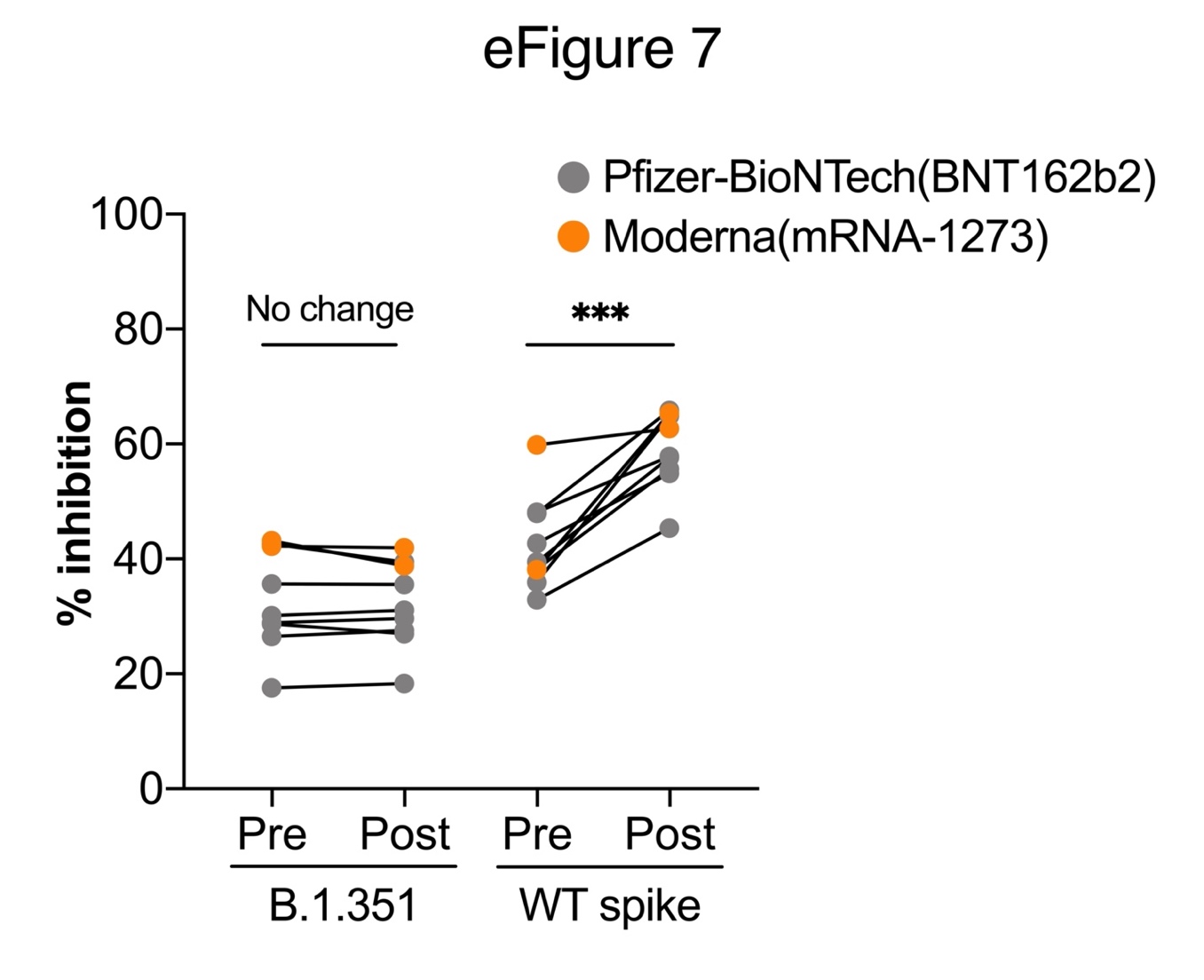
